# Heterogeneity of treatment effect of higher dose dexamethasone by geographic region in patients with COVID-19 and severe hypoxemia - A *post hoc* evaluation of the COVID STEROID 2 trial

**DOI:** 10.1101/2022.11.23.22282463

**Authors:** Bharath Kumar Tirupakuzhi Vijayaraghavan, Anders Granholm, Sheila N Myatra, Vivekanand Jha, Naomi Hammond, Sharon Micallef, Marie Warrer Munch, Maj-Brit N Kjær, Morten Hylander Møller, Theis Lange, Anders Perner, Balasubramanian Venkatesh, the COVID-STEROID 2 collaborators

## Abstract

**Background:** The COVID-STEROID 2 trial found high probability of benefit with dexamethasone 12 mg vs. 6 mg daily among patients with COVID-19 and severe hypoxemia. There was suggestion of heterogeneity of treatment effects (HTE)between patients enrolled from Europe vs. India on the primary outcome. Whether there was HTE by geographical region for the remaining prespecified patient-important outcomes is unclear.

**Methods:** We evaluated HTE by geographical region (Europe vs. India) for all secondary outcomes assessed in the trial with analyses adjusted for stratification variables. The results are presented as risk differences (RDs) or mean differences (MDs) with 99% confidence intervals (CIs) and P-values from interaction tests.

**Results:** We found HTE for mortality at day 28 (RD for Europe -8.3% (99 % CI: -17.7 to 1.0) vs. RD for India 0.1% (99% CI: -10.0 to 10.0)), mortality at day 90 (RD for Europe -7.4% (99% CI: -17.1 to 2.0) vs. RD for India -1.4% (99% CI:-12.8 to 9.8)), mortality at day 180 (RD for Europe -6.7% (99%CI:-16.4 to 2.9) vs. RD for India -1.0% (99%CI:-12.3 to 10.3)), and number of days alive without life support at day 90 (MD for Europe 6.1 days (99% CI:-1.3 to 13.4) vs. MD for India 1.7 days (99% CI:-8.4 to11.8)). For serious adverse reactions, the direction was reversed (RD for Europe -1.0% (99% CI:-7.1 to 5.2) vs. RD for India -5.3% (99% CI: -16.2 to 5.0). For HRQoL outcomes, MD in EQ-5D-5L index values was 0.08(99%CI: -0.01 to 0.16) for Europe and 0.02(99%CI:-0.10 to 0.14) for India. For EQ VAS, MD was 4.4(95%CI:-3.1 to 11.9) for Europe and 2.6(99%CI:-9.0 to 14.2) for India. P values for all tests of interaction were ≥0.12.

**Conclusions:** In this *post hoc* exploratory analysis, we found that higher dose dexamethasone may have lower beneficial effects for patients in India as compared with those in Europe without an increase in serious adverse reactions.

## Background

Clinical trials have demonstrated that corticosteroids as compared to standard care improve survival in patients with COVID-19 needing oxygen and/or advanced forms of respiratory support. (1,2) In the international COVID STEROID 2 randomised trial (n=1000) comparing higher (12 mg) vs lower doses (6 mg) of dexamethasone for patients with COVID-19 and severe hypoxaemia, there was a high probability of benefit from the higher dose for all outcomes assessed up until day 90. (3,4) Long-term outcomes were similarly mostly compatible with benefit (5) Of the analysed trial population, 613 patients were enrolled in Europe (Denmark, Sweden, and Switzerland) and 369 patients in India. There was a suggestion of heterogeneity of treatment effect (HTE) on the primary outcome (days alive without life support at 28 days) when comparing the subgroup of patients enrolled in Europe vs. India [adjusted mean difference (MD) in Europe: 1.8 days (95% confidence interval (CI) 0.2 to 3.4 days) vs India: 0.5 days (95% CI: -1.7 to 2.6)]; however, this was not statistically significant (test of interaction P = 0.57). Potential reasons for HTE between India and Europe include important differences in patient and healthcare system characteristics, resource availability and intensive care capacity, (6,7) the burden of the pandemic, (8) the overall prevalence of healthcare associated infections, (9-11) including the prevalence of infection by multidrug resistant organisms, (12) and concerns of fungal infection outbreaks in India following corticosteroid use. (13,14)

Whether there is HTE according to the geographical regions for the remaining prespecified patient-centred outcomes is unclear. In this *post hoc* exploratory analysis of the COVID-STEROID 2 trial, we assessed whether HTE was present for all the prespecified outcomes for patients enrolled in Europe vs. those enrolled in India. Our hypothesis was that while the overall benefit seen in the full trial population may be preserved, the magnitudes of any benefits are likely lower for the Indian population.

## Methods

This *post hoc* exploratory analysis of HTE in the COVID STEROID 2 trial was conducted according to a prespecified statistical analysis plan (SAP) made available on an online repository. (15) The SAP was written after publication of the original trial results but before any of the analyses reported in this manuscript were conducted.

### The COVID STEROID 2 trial

The COVID STEROID 2 trial was an investigator-initiated, international, parallel-group, blinded randomised clinical trial). (3-5) Detailed descriptions of the trial methods, interventions, outcomes, statistical analyses, and the results for the COVID STEROID 2 trial have been published elsewhere. (3-5) In brief, 1000 adult patients hospitalised with COVID-19 and severe hypoxaemia (requiring ≥10 L oxygen/minute or mechanical ventilation) were enrolled from 31 sites in 26 hospitals in Denmark, India, Sweden and Switzerland between 27 August 2020 and 20 May 2021. Patients were randomised 1:1 to dexamethasone 12 mg or 6 mg intravenously (IV) once daily for up to 10 days.

### Outcomes

Heterogeneity of treatment effects was evaluated for all secondary outcomes assessed in the main trial:

- all-cause mortality at day 28
- number of participants with one or more serious adverse reactions (SARs) from randomisation to day 28 defined as new episodes of septic shock, invasive fungal infection, clinically important gastrointestinal (GI) bleeding or anaphylactic reaction to intravenous dexamethasone
- all-cause mortality at day 90
- days alive without life support at day 90
- days alive and out of hospital at day 90
- all-cause mortality at day 180
- health-related quality of life (HRQoL) at day 180 using EuroQol 5-dimension 5-level (EQ-5D-5L) questionnaire index values and the EuroQol visual analogue scale (EQ VAS)

For the HRQoL outcomes, the defined measures were the EQ-5D-5L index values, i.e., summary scores based on the 5 domains of the EQ-5D-5L questionnaire reflecting the patient’s self-rated health and analysed according to the general population value sets. (16) This ranges from 1.0 (perfect health) to values below zero (health states valued worse than death) with zero defined as a state equivalent to death. (16) The EQ VAS is the patient’s self-rated health ranging from 0 (worst imaginable health) to 100 (best imaginable health). Non-survivors were assigned a value of 0 for both HRQoL outcomes. We used the country specific value sets to calculate the index values for Danish, (17) Indian, (18), and Swedish, (19) patients, and the German value set (20) for those enrolled in Switzerland as no Swiss value set was available.

### Statistical analysis

#### Descriptive data

We present descriptive data for all baseline variables presented in the primary trial report and all outcomes assessed (including descriptive outcome data for the individual EQ-5D-5L domains) stratified by region (Europe vs. India) and treatment group. Categorical data are presented as counts and percentages, and continuous data as medians and interquartile ranges (IQRs).

#### Analyses

Analytical choices, except where otherwise noted, correspond to those of the primary analyses, (3,5) and all analyses were adjusted for the stratification variables (trial site, age below 70 years, and use of invasive mechanical ventilation). Binary outcomes were analysed using logistic regression and G-computation (using 50,000 bootstrap samples) with results presented as adjusted risk differences (RDs) with 99% CIs for each geographical region. The logistic regression models included an interaction between treatment group and geographical region, but no main effect for geographical region as this was already covered by site. (15) Continuous outcomes were analysed separately in each geographical region using linear regression with bootstrapping (50,000 samples) with results presented as adjusted MDs with 99% CIs separately for each geographical region.

For evaluating the HTE, we used tests-of-interactions (Wald’s tests for continuous outcomes and likelihood ratio tests for binary outcomes). P-values for the interaction are presented. Finally, we include Kaplan Meier survival curves (up to day 180) stratified by treatment group and geographical region. As outlined in the analysis plan, results are not dichotomised according to P-value thresholds.

### Handling of missing data

The proportions of missing data for all baseline and outcome variables are presented. Complete case analyses were conducted due to negligible missing data (≤2.0%) for all outcomes except the two HRQoL outcomes. For the HRQoL outcomes, missingness was 6.1% for EQ-5D-5L index values and 5.9% for EQ VAS scores, and thus, multiple imputation was used with all analyses of these outcomes conducted using the multiply imputed datasets only (5,15) We used predictive mean matching with 25 datasets imputed separately in each treatment group. We included the stratification variables, important baseline prognostic variables (age, co-morbidities, use of life support at baseline, geographical region (Europe vs. India), and all outcomes in the imputation model.

#### Software

Analyses were conducted using R (R Core Team, R Foundation for Statistical Computing, Vienna, Austria) v. 4.1.0.

#### Ethics

This was secondary analysis of previously published data and as such did not need a new ethics approval. The original trial was approved by the Institutional Ethics Committee at Apollo Main Hospital (AMH-C-S-021/06-20).

## Results

Descriptive baseline data for the 982 patients in the ITT population are presented in **Table 1** stratified by geographical region and treatment allocation. Baseline characteristics were largely similar between treatment arms within geographical regions. Between geographical regions, baseline characteristics differed for weight (lower in India, prevalence of diabetes (higher in India), time from onset of symptoms to hospitalisation (shorter in India), place of enrolment (almost solely from intensive care units in India), receipt of non-invasive ventilation/continuous positive airway pressure therapy (higher in India), use of invasive mechanical ventilation (lower in India), and use of antivirals and other treatments (higher in India).

**Table 2 and Figures 1 and 2** presents the results of the subgroup analysis for all secondary outcomes. There were differences in several outcomes between geographical regions; mortality at day 28 (adjusted RD for Europe -8.3% (99% CI: -17.7 to 1.0) vs. adjusted RD for India 0.1% (99% CI: - 10 to10)), mortality at day 90 (adjusted RD for Europe -7.4% (99% CI: -17.1 to 2.0) vs. adjusted RD for India -1.4% (99% CI:-12.8 to 9.8)), mortality at day 180 (adjusted RD for Europe -6.7% (99% CI:-16.4 to 2.9) vs. adjusted RD for India -1.0% (99% CI:-12.3 to10.3)), number of days alive without life support at day 90 (adjusted MD for Europe 6.1 days (99% CI:-1.3 to 13.4) vs. adjusted MD for India 1.7 days (99% CI:-8/4 to 11.8)). For SARs, this difference was reversed with a lower proportion of patients in India experiencing SARs from the higher dose (adjusted RD for Europe -1.0% (99% CI:-7.1 to 5.2) vs. adjusted RD for India -5.3% (99% CI: -16.2 to 5.0)). For HRQoL outcomes, the adjusted mean difference in EQ-5D-5L index values between the treatment arms was 0.08(99%CI: -0.01 to 0.16) for Europe as compared to 0.02(99%CI:-0.10 to 0.14) for India. For EQ VAS, the adjusted mean difference was 4.4(95%CI:-3.1 to 11.9) for Europe as compared to 2.6(99%CI:-9.0 to 14.2) for India. P values for all tests of interaction were ≥0.12. **(Table 2 and Supplementary Table 1)**

**Figure 1:**
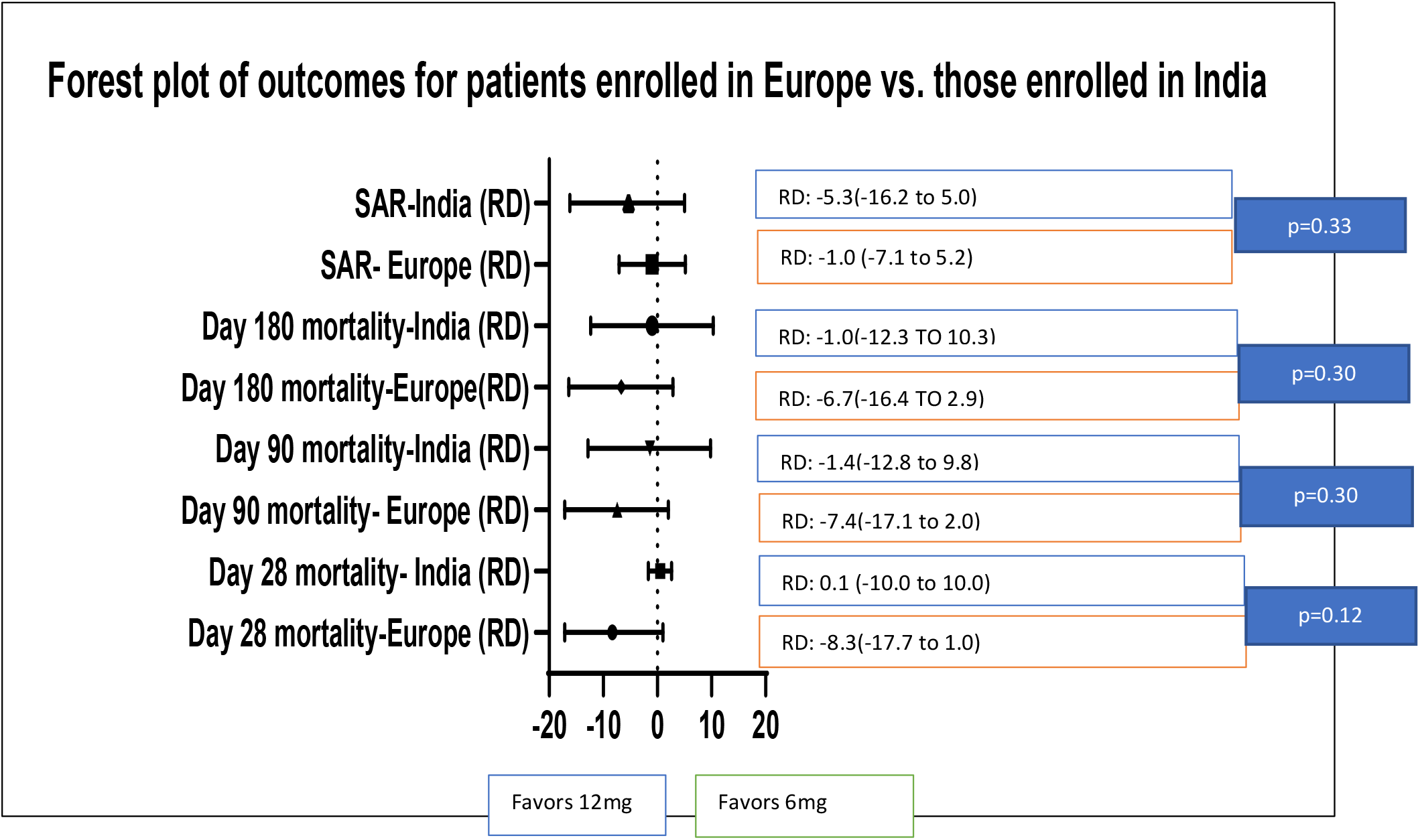
Forest plot of categorical outcomes between Europe and India for the 12mg vs. 6mg Dexamethasone comparison.

**Figure 2:**
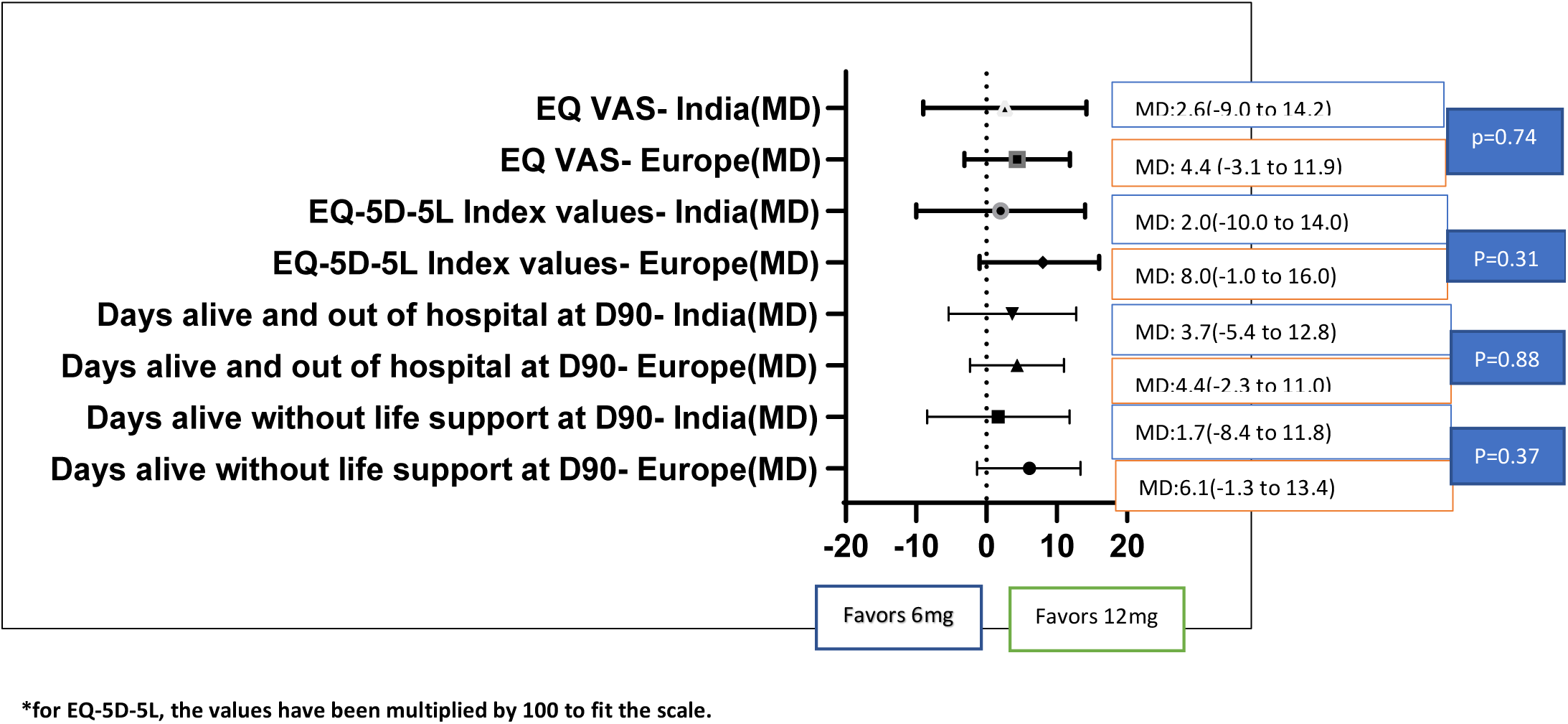
Forest plot of continuous outcomes between Europe and India for the Dexamethasone 12mg versus 6mg comparison.

**Figure 3**. presents the Kaplan Meier survival curve by treatment arm and by region illustrating the higher beneficial effect of 12 mg for patients enrolled in Europe compared with India.

**Figure 3:**
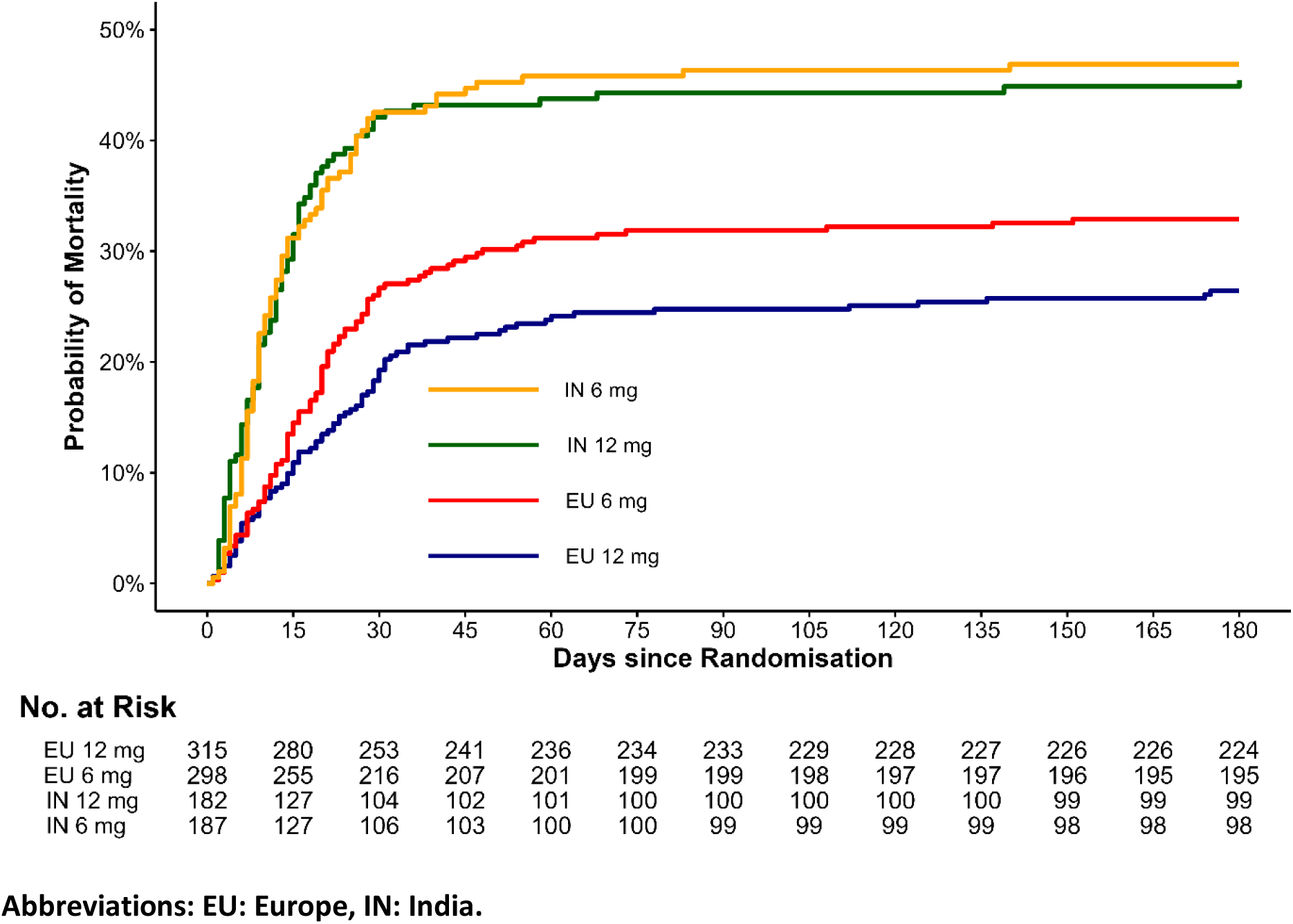
Morality at Day 180 - Europe vs. India. **Abbreviations: EU: Europe, IN: India.**

## Discussion

In this *post hoc* exploratory analysis, we found that any benefits of 12 mg vs. 6 mg dexamethasone appeared to be reduced for patients enrolled in India for a number of outcomes.-day 28, day 90 and day 180 mortality and fewer number of days alive without life support at day 90. Reassuringly, there did not appear to be an increase in the occurrence of SARs at day 28 in patients enrolled in India.

Exploring treatment subgroup effects for patients enrolled from different health system contexts is valuable for a number of reasons; including important differences in patient and healthcare system characteristics, baseline risk factors, resource availability, and intensive care capacity. (6-8) For treatments such as higher doses of corticosteroids, additional factors limited to lower-middle income countries (LMICs) may also play a role such as the higher potential for harm due to the differing comorbidity burden, and the higher prevalence of both healthcare associated infections including infections caused by multidrug resistant organisms. (9-13) In this *post hoc* analysis, the differences in baseline characteristics such as the higher prevalence of diabetes and the higher proportion of patients being enrolled from ICUs in India may have contributed to the heterogeneity of effects.

Interestingly, such heterogeneity of treatment effect by geographical region has previously been demonstrated in the context of corticosteroid use in bacterial meningitis, specifically caused by *Streptococcus pneumoniae*, in which corticosteroids have been shown to be beneficial in high income countries (relative risk for reduction in mortality: 0.48 (95%CI: 0.24 to 0.96)), (21), while such benefit was not demonstrated in a similar study from Sub-Saharan Africa (odds ratio for mortality: 1.10 (95%CI: 0.68 to1.77)). (22) Much of the postulated reasons for these findings were related to the differing comorbid profile (e.g., a higher human immunodeficiency virus (HIV) prevalence in Africa), challenges in accessing healthcare, and differences in pathogen profiles among others. Specific to India, concerns have been raised over the risk of secondary sepsis and fungal infections contributing to worse outcomes. (13,14,23) This assumes greater importance given the higher prevalence of diabetes mellitus in India (11.8% in India vs. 6.2% in European countries) (24,25)and the consequent higher risk of such infections. In the primary analyses of the COVID STEROID 2 trial, (3) we found no major differences in the incidence of new episodes of septic shock or invasive fungal infections between the treatment arms. While we do not have information on this outcome beyond day 28, this subgroup analysis provides further reassurance about the safety of the higher dose in patients from India.

The lower benefit seen for patients enrolled in India with the 12 mg dose may be driven by the differences in baseline characteristics. Although, we had hypothesised that one of the reasons for this difference might be a delayed presentation to the hospital, median time from symptom onset to hospitalisation was 5 days in India as compared to 8 days in Europe. Finally, racial and ethnic differences in the effects of inhaled corticosteroid use on bronchodilator response have been reported in patients with asthma. (26) Whether similar factors may play a role in the response to parenteral corticosteroids in patients with COVID-19 is unknown.

### Strengths and limitations

The strengths of the study include the strengths of the COVID STEROID 2 trial, i.e., a large, international, blinded randomised controlled trial with high rates of follow-up. Importantly, this analysis provides additional information and context for clinicians working in both Europe (and other high-income countries) and India (and other similar LMICs) on the effects and safety profile of higher doses of corticosteroids for patients with COVID-19 and severe hypoxaemia.

Our study has limitations too, including those general to the COVID STEROID 2 trial, i.e., the evolving pandemic and changes in care during and after the trial, e.g., recommendations in favour of interleukin-6 receptor antagonists introduced after randomisation concluded, (27) and limited power for some outcomes and analyses, including these secondary subgroup analyses. Moreover, we collected information on new episodes of septic shock or invasive fungal infection only up to day 28. Longer-term information on this outcome would be of interest to clinicians in India and other LMICs. Finally, this was a *post hoc* exploratory study, and despite public registration of the statistical analysis plan prior to conduct of the analyses, this was done after the primary trial results were known. Consequently, these results should be interpreted cautiously and as hypothesis-generating only.

## Conclusions

In this *post hoc* exploratory analysis, we found that higher dose dexamethasone may have lower beneficial effects for patients with COVID-19 and severe hypoxaemia in India as compared to those in Europe without an increase in serious adverse reactions.

## Data Availability

Data are available upon reasonable request. Deidentified patient data will be made available beginning 6 months after publication of the study and ending at 2 years. All requests for data sharing must be accompanied by a formal request, a study proposal with clear statement of aims and hypotheses and a statistical analysis plan. All applications will be assessed by the COVID-STEROID 2 Trial Management Committee. Applications from investigators with suitable academic capability to conduct the proposed work will be given consideration. Any proposal will require approval from the ethics committee which approved the conduct of this trial prior to sharing of any patient data. If a proposal is approved, a signed data transfer agreement will be required before data sharing.

## References

1. The RECOVERY Collaborative Group. Dexamethasone in Hospitalized Patients with COVID-19. N Engl J Med 2021;384:693–704

2. The WHO Rapid Evidence Appraisal for COVID-19 Therapies (REACT) Working Group. Association Between Administration of Systemic Corticosteroids and Mortality Among Critically Ill Patients with COVID-19: A Meta-analysis. JAMA

3. The COVID STEROID 2 Trial group. Effect of 12mg vs. 6mg of Dexamethasone on the Number of Days Alive Without Life Support in Adults with COVID-19 and Severe Hypoxemia. JAMA 2021;326(18):1807–17

4. Granholm A, Munch MW, Myatra SN, Tirupakuzhi Vijayaraghavan BK, Cronhjort M, Wahlin RR et al. Dexamethasone 12mg versus 6mg for patients with COVID-19 and severe hypoxaemia: a pre-planned, secondary Bayesian analysis of the COVID-STEROID 2 trial. Intensive Care Med 2022;48(1):45–55

5. Granholm A, Kjaer MN, Wunch MW, Myatra SN, Tirupakuzhi Vijayaraghavan BK, Cronhjort M et al. Long-term outcomes of dexamethasone 12mg versus 6mg in patients with COVID-19 and severe hypoxaemia. Intensive Care Med 2022;48(5):580–89

6. Phua J, Faruq MO, Kulkarni AP, Redjeki IS, Detleuxay K, Mendsaikhan N et al. Critical Care Bed Capacity in Asian Countries and Regions. Crit Care Med 2020;48(5)654–62

7. Tirupakuzhi Vijayaraghavan BK, Myatra SN, Mathew M, Lodh N, Divatia JV, Hammond N et al. Challenges in the delivery of critical care in India during the COVID-19 pandemic. J Intensive Care Soc 2021;22(4):342–48

8. Available from: https://covid19.who.int/region/searo/country/in (accessed on 19th October 2022)

9. Rosenthal VD, Bat-Erdene I, Gupta D, Belkebir S, Rajhans P, Zand F et al. International Nosocomial Infection Control Consortium (INICC) report, data summary of 45 countries for 2012-17: Device-associated module. Am J Infect Control 2020;48(4):423–32

10. Mathur P, Malpiedi P, Walia K, Srikantiah P, Gupta S, Lohiya A et al. Health-care-associated bloodstream and urinary tract infections in a network of hospitals in India: a multicentre, hospital-based prospective surveillance study. Lancet Glob Health 2022;10(9):e1317–25

11. Murhekar MV, Girish Kumar CP. Health-care-associated infection surveillance in India. Lancet Glob Health 2022;10(9):e1222–23

12. Venkataraman R, Divatia JV, Ramakrishnan N, Chawla R, Amin P, Gopal P et al. Multicenter Observational Study to Evaluate Epidemiology and Resistance Patterns of Common Intensive Care Unit-infections. Indian J Crit Care Med 2018;22(1):20–26

13. Available from: https://www.cdc.gov/fungal/covid-fungal.html (accessed on 19th October 2022)

14. Hoenigl M, Seidel D, Sprute R, Cunha C, Oliverio M, Goldman GH et al. COVID-19 associated fungal infections. Nature Microbiology 2022;7:1127–40

15. Available from: https://osf.io/6tfrk (accessed on 19th October 2022)

16. Herdman M, Gudex C, Lloyd A, Janssen M, Kind P, Parkin D, Bonsel G, Badia X (2011) Development and preliminary testing of the new five-level version of EQ-5D (EQ-5D-5L). Qual Life Res 20:1727–1736

17. Jensen CE, Sorenson SS, Gudex C et al. The Danish EQ-5D-5L value set: a hybrid model using cTTO and DCE data. Appl Health Econ Health Policy 2021;19:579–91

18. Jyani G, Prinja S, Kar SS et al. Valuing health-related quality of life among the Indian population: a protocol for the development of an EQ-5D value set for India using an extended design (DEVINE) Study. BMJ Open 2020; 10:e039517

19. Burstrom K, Teni FS, Gerdtham UG et al. Experience-based Swedish TTO and VAS value sets for EQ-5D-5L health states. Pharmacoeconomics 2020;38:839–56

20. Ludwig K, Graf von der Schulenburg JM, Griener W et al. German Value Set for the EQ-5D-5L. Pharmacoeconomics 2018;36:663–674

21. Gans dJ, van de Beek D. Dexamethasone in Adults with Bacterial Meningitis. N Engl J Med 2002;347

22. Scarborough M, Gordon SB, Whitty CJM, French N, Njalale Y, Chitani A et al. Corticosteroids for Bacterial Meningitis in Adults in Sub-Saharan Africa. N Engl J Med 2007;357:2441–50

23. Singh AK, Singh R, Joshi SR, Misra A. Mucormycosis in COVID-19: A systematic review of cases reported worldwide and in India. Diabetes Metab Syndr 2021;15(4):102146

24. Available from: https://www.livemint.com/science/health/government-survey-found-11-8-prevalence-of-diabetes-in-india-11570702665713.html (accessed on 3rd November 2022)

25. Available from: https://www.oecd-ilibrary.org/sites/83231356-en/index.html?itemId=/content/component/83231356-en (accessed on 3rd November 2022)

26. Samedy-Bates LA, Oh SS, Nuckton TJ, Elhawary JR, White M, Elliot T et al. Racial/Ethnic - Specific Differences in the Effects of Inhaled Corticosteroid Use on Bronchodilator Response in Patients with Asthma. Clin Pharmacol Ther 2019;106(5):1133–40

27. Available from: http://surl.li/djzfx (accessed on 19th October 2022)

