## Supplementary material for "Heterogeneity of treatment effect of higher dose dexamethasone by geographic region in patients with COVID-19 and severe hypoxemia - A *post hoc* evaluation of the COVID STEROID 2 trial": Tables

**Table 1: Baseline characteristics**

| **Baseline variable** | **Europe** | | **India** | |
| --- | --- | --- | --- | --- |
|  | 12mg (n=315) | 6mg (n=298) | 12mg (n=182) | 6mg (n=187) |
| **Country of Inclusion** |  |  |  |  |
| - Denmark | 251 (79.7%) | 234 (78.5%) | - | - |
| - Sweden | 40 (12.7%) | 39 (13.1%) | - | - |
| - Switzerland | 24 (7.6%) | 25 (8.4%) | - | - |
| - India | - | - | 182 (100%) | 187 (100%) |
| Age in years  - median (IQR) | 65.0  (57.0-74.5) | 66.0  (57.0-73.8) | 63.5  (54.0-70.0) | 61.0  (51.5-70.0) |
| Sex (n and %)   - Male | 214 (67.9%) | 214 (71.8%) | 132 (72.5%) | 117 (62.6%) |
| Weight in kgs  - median (IQR) | 90.0  (76.0-104.5) | 90.0  (77.0-100.0) | 65.5 (60.0-73.8) | 68.0 (60.0-75.0) |
| **Coexisting conditions (n and %)** |  |  |  |  |
| - Ischemic heart disease or heart failure | 46 (14.6%) | 47 (15.8%) | 21 (11.5%) | 22 (11.8%) |
| - Diabetes mellitus | 62 (19.7%) | 87 (29.2%) | 73 (40.1%) | 76 (40.6%) |
| - Chronic obstructive pulmonary disease | 49 (15.6%) | 46 (15.4%) | 8 (4.4%) | 10 (5.3%) |
| - Immunosuppressive therapy within 3 months prior to randomization | 27 (8.6%) | 24 (8.1%) | 13 (7.1%) | 19 (10.2%) |
| - Chronic use of systemic glucocorticoids | 11 (3.5%) | 15 (5.0%) | 2 (1.1%) | 1 (0.5%) |
| Limitations in the use of CPR or life support at randomization (n and %) | 29 (9.2%) | 25 (8.4%) | 1 (0.5%) | 1 (0.5%) |
| Time from onset of symptoms to hospitalization in days  - median (IQR)* | 8.0 (5.0-10.0) | 8.0 (5.0-10.0) | 5.0 (3.0-7.0) | 5.0 (3.0-7.0) |
| Time from hospitalization to randomization in days - median (IQR) | 2.0 (1.0-3.0) | 1.0 (1.0-3.0) | 2.0 (1.0-3.0) | 2.0 (1.0-3.0) |
| **Place of enrollment, n and %** |  |  |  |  |
| - Intensive care unit | 222 (70.5%) | 215 (72.1%) | 167 (91.8%) | 178 (95.2%) |
| - Hospital ward | 61 (19.4%) | 53 (17.8%) | 5 (2.7%) | 1 (0.5%) |
| - Emergency department | 14 (4.4%) | 13 (4.4%) | 8 (4.4%) | 8 (4.3%) |
| - Intermediate care unit | 18 (5.7%) | 17 (5.7%) | 2 (1.1%) | 0 (0.0%) |
| **Type of oxygen supplementation** |  |  |  |  |
| **Nasal cannula or open mask,  n and %** | 190 (60.3%) | 176 (59.1%) | 82 (45.1%) | 82 (43.9%) |
| *Flow rate in L/min,  median (IQR)* | 23.5 (15.0-37.0) | 25.0 (15.0-40.0) | 15.0 (12.0-45.0) | 16.0 (12.0-48.8) |
| **Noninvasive ventilation or continuous positive airway pressure, n and %** | 39 (12.4%) | 45 (15.1%) | 79 (43.4%) | 83 (44.4%) |
| *Fio2 in %, median (IQR)^#^* | 80.0 (67.5-99.0) | 70.0 (64.0-100.0) | 50.0  (50.0-60.0) | 50.0 (40.0-60.0) |
| *Duration before randomization in days, median (IQR)* | 1.0  (0.0-1.0) | 1.0  (0.0-1.0) | 1.0  (0.0-1.0) | 1.0  (0.0-1.5) |
| **Invasive ventilation, n and %** | 86 (27.3%) | 77 (25.8%) | 21 (11.5%) | 22 (11.8%) |
| *Fio2 in %, median (IQR)^$^* | 55.0  (45.0-70.0) | 60.0 (45.0-90.0) | 70.0 (60.0-90.0) | 62.5 (50.0-70.0) |
| *Duration before randomization in days, median (IQR)* | 1.0  (0.0-1.0) | 1.0  (0.0-1.0) | 1.0  (0.0-1.0) | 1.0  (1.0-1.0) |
| **Baseline Oxygenation status** |  |  |  |  |
| *PaO2 in mmHg (median/IQR)^* | 69.8 (61.5-83.1) | 70.0 (62.2-81.0) | 76.5 (60.8-100.0) | 72.0 (58.8-83.4) |
| *Saturation in % (median/IQR)^&^* | 94.0 (91.0-96.0) | 93.0 (91.0-96.0) | 95.0 (91.0-97.0) | 94.0 (90.0-96.5) |
| Lactate concentration in mg/dl (median/IQR)^@^ | 15.3 (11.7-23.4) | 15.8 (11.7-21.6) | 10.8 (6.3-18.0) | 13.5 (8.1-18.2) |
| Vasopressors or inotropes | 68 (21.6%) | 57 (19.1%) | 13 (7.1%) | 11 (5.9%) |
| Kidney replacement therapy | 7 (2.2%) | 7 (2.3%) | 4 (2.2%) | 7 (3.7%) |
| **Anti-inflammatory agents** | 31 (9.8%) | 31 (10.4%) | 27 (14.8%) | 26 (13.9%) |
| *IL-6 receptor antagonists* | 25(7.9%) | 23 (7.7%) | 27 (14.8%) | 24 (12.8%) |
| *Janus kinase inhibitor* | 0 (0.0%) | 0 (0.0%) | 8 (4.4%) | 7 (3.7%) |
| *Other* | 7 (2.2%) | 8 (2.7%) | 2 (1.1%) | 2 (1.1%) |
| **Antiviral agents** | 162(51.4%) | 157(52.7%) | 150(82.4%) | 161(86.1%) |
| *Remdesivir* | 157(49.8%) | 151 (50.7%) | 150 (82.4%) | 159 (85.0%) |
| *Convalescent plasma* | 4(1.3%) | 7(2.3%) | 7(3.8%) | 10(5.3%) |
| *Other* | 9(2.9%) | 4(1.3%) | 0(0.0%) | 2(1.1%) |

* proportion missing 5.1%; # missing 1.2% ; $ missing 0.1%; ^missing 5.2%; & missing 1.4%; @missing 10.8%

Add all abbreviations

**Table 2: Outcomes**

| **Outcome** | **Europe** | | **Effect estimates with 99% CI^*^** | **India** | | **Effect estimates with 99% CI** | **P value for test of interaction** |
| --- | --- | --- | --- | --- | --- | --- | --- |
|  | 12mg (n=315) | 6mg (n=298) |  | 12mg (n=182) | 6mg (n=187) |  |  |
| Mortality at day 28-n (%) | 57 (18.3%) | 76 (25.8%) | -8.3 (-17.7 to 1) | 76 (42.2%) | 79 (42.7%) | 0.1 (-10.0 to 10.0) | 0.12 |
| Serious adverse reactions at day 28 - n (%) | 40 (12.7%) | 41 (13.8%) | -1.0 (-7.1 to 5.2) | 16 (8.8%) | 24 (12.8%) | -5.3 (-16.2 to 5) | 0.33 |
| Mortality at day 90 - n (%) | 77 (24.8%) | 94 (32.1%) | -7.4 (-17.1 to 2.0) | 80 (44.4%) | 86 (46.5%) | -1.4 (-12.8 to 9.8) | 0.30 |
| No. of days alive without life support at day 90- median (IQR) | 83.0 (35.5-90.0) | 80.0 (8.0-90.0) | 6.1 (-1.3 to 13.4) | 89.5 (5.0-90.0) | 74.5 (5.0-90.0) | 1.7 (-8.4 to 11.8) | 0.37 |
| No. of days alive and out of hospital at day 90- median (IQR) | 63.0 (0.0-77.8) | 54.0 (0.0-76.0) | 4.4 (-2.3 to 11.0) | 50.5 (0.0-78.0) | 0.0 (0.0-78.0) | 3.7 (-5.4 to 12.8) | 0.88 |
| Mortality at day 180- n (%) | 82  (26.8%) | 97 (33.2%) | -6.7 (-16.4 to 2.9) | 82 (45.6%) | 87 (47.0%) | -1.0 (-12.3 to 10.3) | 0.30 |
| EQ-5D-5L index values- median (IQR)^$^ | 0.80 (0.00-0.92) | 0.67 (0.00-0.91) | 0.08 (-0.01 to 0.16) | 0.85 (0.00-1.00) | 0.74 (0.00-1.00) | 0.02 (-0.10 to 0.14) | 0.31 |
| EQ VAS – median (IQR)^$^ | 60.0 (0.0-80.0) | 55.0 (0.0-80.0) | 4.4 (-3.1 to 11.9) | 80.0 (0.0-100.0) | 65.0 (0.0-95.0) | 2.6 (-9.0 to 14.2) | 0.74 |

***For continuous outcomes, adjusted mean differences (in days) and for binary outcomes, adjusted risk differences (in percentage points) are presented. Adjustment is for stratification variables.**

**$These analyses are based on imputed datasets for missing values. All others are complete case analyses.**
