## Supplementary material for "Heterogeneity of treatment effect of higher dose dexamethasone by geographic region in patients with COVID-19 and severe hypoxemia - A *post hoc* evaluation of the COVID STEROID 2 trial": suppl.material

**Supplementary Table 1:** Number (%) of respondents for each level within the components of the EQ5D5L amongst survivors at 180 days

| **EQ-5D-5L Component** | **Europe** | | **India** | |
| --- | --- | --- | --- | --- |
|  | 12 mg (n=224) | 6 mg (n=195) | 12 mg (n=98) | 6 mg  (n=98) |
| **Mobility** |  |  |  |  |
| - 1 (no problems) | 90 (44.6%) | 73 (40.6%) | 90 (91.8%) | 82 (85.4%) |
| - 2 (slight problems) | 53 (26.2%) | 43 (23.9%) | 6 (6.1%) | 10 (10.4%) |
| - 3 (moderate problems) | 39 (19.3%) | 37 (20.6%) | 1 (1.0%) | 4 (4.2%) |
| - 4 (severe problems) | 16 (7.9%) | 22 (12.2%) | 0 (0.0%) | 0 (0.0%) |
| - 5 (unable to walk around) | 4 (2.0%) | 5 (2.8%) | 1 (1.0%) | 0 (0.0%) |
| **Self-care** |  |  |  |  |
| - 1 (no problems with washing or dressing) | 159 (78.7%) | 144 (80.0%) | 89 (90.8%) | 85 (88.5%) |
| - 2 (slight problems) | 23 (11.4%) | 19 (10.6%) | 8 (8.2%) | 6 (6.2%) |
| - 3 (moderate problems) | 11 (5.4%) | 12 (6.7%) | 0 (0.0%) | 5 (5.2%) |
| - 4 (severe problems) | 2 (1.0%) | 3 (1.7%) | 0 (0.0%) | 0 (0.0%) |
| - 5 (unable to wash or dress) | 7 (3.5%) | 2 (1.1%) | 1 (1.0%) | 0 (0.0%) |
| **Usual activities** |  |  |  |  |
| - 1 (no problems with usual activities) | 65 (32.3%) | 68 (37.8%) | 92 (93.9%) | 83 (86.5%) |
| - 2 (slight problems) | 65 (32.3%) | 47 (26.1%) | 4 (4.1%) | 8 (8.3%) |
| - 3 (moderate problems) | 44 (21.9%) | 47 (26.1%) | 1 (1.0%) | 5 (5.2%) |
| - 4 (severe problems) | 13 (6.5%) | 16 (8.9%) | 0 (0.0%) | 0 (0.0%) |
| - 5 (unable to perform) | 14 (7.0%) | 2 (1.1%) | 1 (1.0%) | 0 (0.0%) |
| **Pain/discomfort** |  |  |  |  |
| - 1 (no pain or discomfort) | 84 (41.6%) | 73 (40.6%) | 84 (85.7%) | 71 (74.0%) |
| - 2 (slight) | 58 (28.7%) | 52 (28,9%) | 10 (10.2%) | 24 (25.0%) |
| - 3 (moderate) | 43 (21.3%) | 33 (18,3%) | 2 (2.0%) | 0 (0.0%) |
| - 4 (severe) | 16 (7.9%) | 19 (10.6%) | 1 (1.0%) | 1 (1.0%) |
| - 5 (extreme pain or discomfort) | 1 (0.5%) | 3 (1.7%) | 1 (1.0%) | 0 (0.0%) |
| **Anxiety/depression** |  |  |  |  |
| - 1 (no anxiety or depression) | 130 (64.7%) | 98 (54.4%) | 86 (87.8%) | 81 (84.4%) |
| - 2 (slight) | 42 (20.9%) | 46 (25.6%) | 10 (10.2%) | 10 (10.4%) |
| - 3 (moderate) | 23 (11.4%) | 25 (13.9%) | 1 (1.0%) | 3 (3.1%) |
| - 4 (severe) | 4 (2.0%) | 9 (5.0%) | 0 (0.0%) | 2 (2.1%) |
| - 5 (extremely anxious or depressed | 2 (1.0%) | 2 (1.1%) | 1 (1.0%) | 0 (0.0%) |

**Full list of COVID-STEROID 2 Collaborators**

| Marie W. Munch MD; | [](about:blank) |
| --- | --- |
| Sheila N. Myatra PhD; | [](about:blank) |
| Bharath Kumar Tirupakuzhi Vijayaraghavan MD; | [](about:blank) |
| Sanjith Saseedharan MD, | [](about:blank) |
| Thomas Benfield PhD; | [](about:blank) |
| Rebecka R. Wahlin PhD; | [](about:blank) |
| Bodil S. Rasmussen PhD; | [](about:blank) |
| Anne Sofie Andreasen PhD; | [](about:blank) |
| Lone M. Poulsen MD, | [](about:blank) |
| Luca Cioccari MD; | [](about:blank) |
| Mohd S. Khan MD; | [](about:blank) |
| Farhad Kapadia MD; | [](about:blank) |
| Jigeeshu V. Divatia MD; | [](about:blank) |
| Anne C. Brøchner PhD; | [](about:blank) |
| Morten H. Bestle PhD; | [](about:blank) |
| Marie Helleberg PhD; | [](about:blank) |
| Jens Michelsen PhD; | [](about:blank) |
| Ajay Padmanaban MD; | [](about:blank) |
| Neeta Bose MD; | [](about:blank) |
| Anders Møller PhD; | [](about:blank) |
| Kapil Borawake MD; | [](about:blank) |
| Klaus T. Kristiansen MD; | [](about:blank) |
| Urvi Shukla MD; | [](about:blank) |
| Michelle S. Chew PhD; | [](about:blank) |
| Subhal Dixit MD; | [](about:blank) |
| Charlotte S. Ulrik PhD; | [](about:blank) |
| Pravin R. Amin MD; | [](about:blank) |
| Rajesh Chawla MD; | [](about:blank) |
| Christian A. Wamberg MD; | [](about:blank) |
| Mehul S. Shah MD; | [](about:blank) |
| Iben S. Darfelt MD; | [](about:blank) |
| Vibeke L. Jørgensen PhD; | [](about:blank) |
| Margit Smitt MD; | [](about:blank) |
| Anders Granholm MD; | [](about:blank) |
| Maj-Brit N. Kjær MSc (Health); | [](about:blank) |
| Morten H. Møller PhD; | [](about:blank) |
| Tine S. Meyhoff MD; | [](about:blank) |
| Gitte K. Vesterlund MSc (Health); | [](about:blank) |
| Naomi E. Hammond PhD;* | [](about:blank) |
| Sharon Micallef BN; | [](about:blank) |
| Abhinav Bassi PT; | [](about:blank) |
| Oommen John MD, MBA;* | [](about:blank) |
| Anubhuti Jha MD; | [](about:blank) |
| Maria Cronhjort PhD; | [](about:blank) |
| Stephan M. Jakob PhD; | [](about:blank) |
| Christian Gluud Dr Med Sc; | [](about:blank) |
| Theis Lange PhD; | [](about:blank) |
| Vaijayanti Kadam MD; | [](about:blank) |
| Klaus V. Marcussen MD; | [](about:blank) |
| Jacob Hollenberg PhD; | [](about:blank) |
| Anders Hedman MD; | [](about:blank) |
| Henrik Nielsen DMSci; | [](about:blank) |
| Olav L. Schjørring PhD; | [](about:blank) |
| Marie Q. Jensen BSc; | [](about:blank) |
| Jens W. Leistner BSc; | [](about:blank) |
| Trine B. Jonassen BSC; | [](about:blank) |
| Camilla M. Kristensen BSc; | [](about:blank) |
| Esben C. Clapp BSc; | [](about:blank) |
| Carl J. S. Hjortsø BSc; | [](about:blank) |
| Thomas S. Jensen MD; | [](about:blank) |
| Liv S. Halstad BSc; | [](about:blank) |
| Emilie R. B. Bak BSc; | [](about:blank) |
| Reem Zaabalawi BSc; | [](about:blank) |
| Matias Metcalf-Clausen BSc; | [](about:blank) |
| Suhayb Abdi BSc; | [](about:blank) |
| Emma V. Hatley BSc; | [](about:blank) |
| Tobias S. Aksnes BSc; | [](about:blank) |
| Emil Gleipner-Andersen BSc; | [](about:blank) |
| A. Felix Alarcón BSc; | [](about:blank) |
| Gabriel Yamin BSc; | [](about:blank) |
| Adam Heymowski BSc; | [](about:blank) |
| Anton Berggren BSc; | [](about:blank) |
| Kirstine la Cour BSc; | [](about:blank) |
| Sarah Weihe BSc; | [](about:blank) |
| Alison H. Pind BSc; | [](about:blank) |
| Janus Engstrøm BSc; | [](about:blank) |
| Vivekanand Jha PhD;* | [](about:blank) |
| Balasubramanian Venkatesh PhD;* | [](about:blank) |
| Anders Perner PhD | [](about:blank) |
